# Rare genetic variants impact muscle strength

**DOI:** 10.1101/2022.06.22.22276715

**Authors:** Yunfeng Huang, Dora Bodnar, Chia-Yen Chen, Gabi Gurria, Mark Sanderson, Biogen Biobank Team, Jun Shi, Katherine G Meilleur, Matthew E. Hurles, Sebastian S. Gerety, Ellen A. Tsai, Heiko Runz

## Abstract

Muscle strength is highly heritable and predictive for multiple adverse health outcomes including mortality. Here, we present a rare protein-coding variant association study in 340,319 individuals for hand grip strength, a proxy measure of muscle strength. We identify six significant genes (*KDM5B, OBSCN, GIGYF1, TTN, RB1CC1, EIF3J*), propose shared mechanisms between brain and muscle function and demonstrate additive effects between rare and common genetic variation on muscle strength.

## Main

Muscle strength is a key measure of physical ability and overall health and when reduced is associated with adverse health outcomes including disability and mortality^1^. Muscle strength is highly heritable (h^2^∼0.4)^2^, but the underlying genetic architecture and mechanisms remain unclear. Hand grip strength (HGS) is a reliable proxy measure of general muscle strength with genome-wide associations studies (GWAS) having identified >180 common-variant based HGS loci^3^. No study has yet systematically analyzed the contribution of rare protein-coding variation to HGS. Here, we leverage whole exome sequencing in 340,319 UK Biobank (UKB)^4^ participants to uncover genes in which rare coding variants are associated with HGS.

We first assessed globally the impact of rare coding variant burden on HGS. Fifteen burden tests were conducted based on predicted effects of rare coding variants (protein-truncating, missense or synonymous), and further on deleteriousness score for missense variants (Fig.1a, TableS1). When stratified by pLI (probability of loss-of-function (LoF) intolerance)^5^, the strongest effect on HGS was observed for PTV-burden from LoF-intolerant genes (pLI>=0.9, beta=-0.25 kg, p=4.05e-56). Significant associations (p<0.05/15=0.0033) with reduced HGS were further observed for PTV-burden in LoF-tolerant genes, very damaging and damaging missense variant burden.

**Figure 1.**
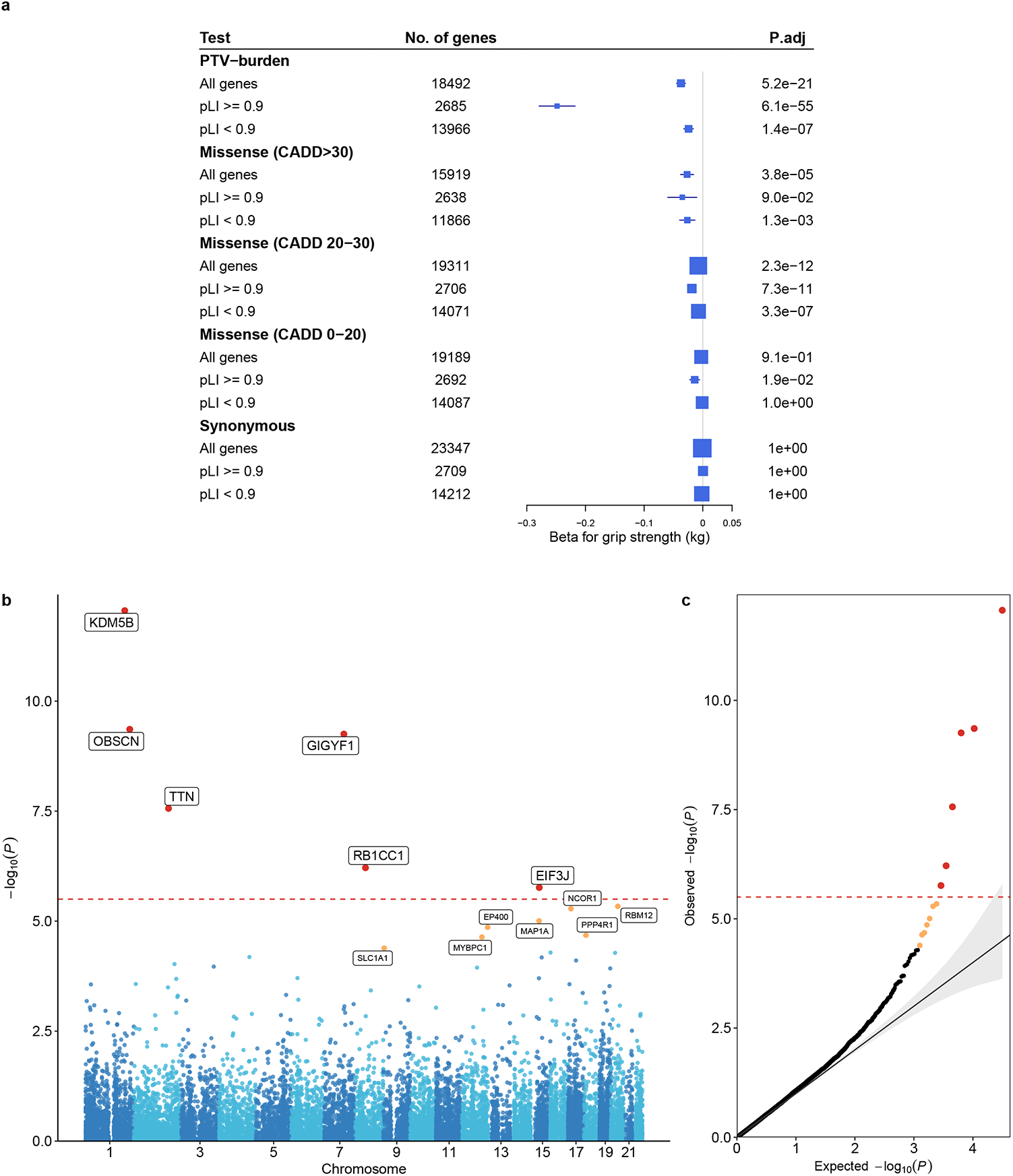
Genome-wide rare variant burden associations with HGS. **a)** Rare variant burden associations with HGS stratified on PTV, very damaging missense (CADD>30), damaging missense (CADD 20-30), other missense (CADD 0-20) and synonymous variants. Associations were tested for all, LoF intolerant (pLI>0.9) and LoF tolerant (pLI<0.9) genes, respectively. **b-c)** Manhattan and QQ plot of gene-based PTV-burden associations with HGS. Red dashed line indicates Bonferroni significance threshold, red circle indicates significant genes that passed the Bonferroni threshold, orange circles indicate genes with FDR<0.05.

Next, we conducted gene-level association analyses to identify individual genes in which a burden of PTV or missense variants affects HGS. For 6 genes (*KDM5B, OBSCN, GIGYF1, TTN, RB1CC1, EIF3J*), PTV-burden showed significant association with HGS after Bonferroni correction (p<3.2e-6) (Fig.1b, TableS2-4).

Notably, two HGS genes identified were giant sarcomeric proteins titin (*TTN*) and obscurin (*OBSCN*), which are essential in maintaining structure and function of striated muscle, with mutations in *TTN* causing Mendelian diseases affecting skeletal and cardiac muscle (OMIM #188840). 3,925 *TTN* PTV-carriers in UKB showed an average of -0.61 kg reduction of HGS relative to non-carriers (p=2.7e-8, Fig.2a). *TTN* PTV-burden was also associated with risk of cardiomyopathy, atrial fibrillation and reduced muscle mass (Fig.S1, TableS5; for findings on other HGS genes, see Supplementary Information).

**Figure 2.**
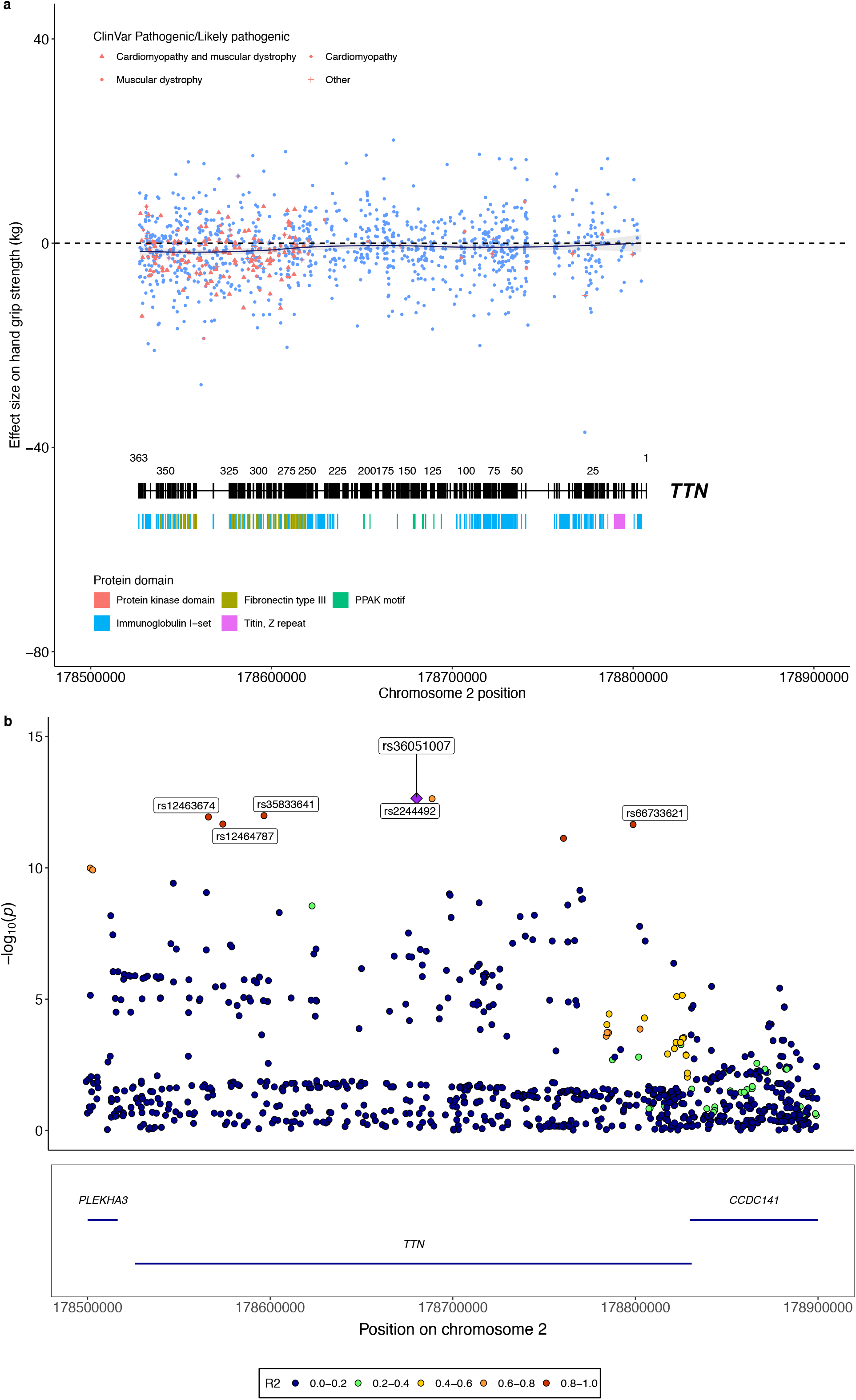
Converging rare and common variant associations of *TTN* with HGS. **a)** Single PTV associations of *TTN* with HGS. Average effect sizes of each PTV in *TTN* were plotted against their genomic positions, with exon numbers and protein domains visualized at the bottom. ClinVar pathogenic/likely pathogenic PTVs were colored in red with different shapes indicating their respective associated disease phenotypes. **b)** HGS GWAS locus plot of *TTN*. Purple diamond indicates the sentinel variant at the *TTN* GWAS locus. SNP IDs label the top 95% credible set identified by SuSiE. Colors indicate different levels of LD with the sentinel variant.

We next assessed whether also common variants (MAF>0.01) in *TTN* were associated with HGS by conducting a GWAS in UKB followed by a gene-based analysis using MAGMA (v1.6)^6^. Indeed, *TTN* showed a significant gene-based association (p=5.5e-8) revealing that both rare and common genetic variants in *TTN* impact muscle strength. We utilized the rare PTV association results of *TTN* to fine-map the *TTN* locus. Statistical fine-mapping identified a top 95% credible set of six variants including the damaging missense variant rs12463674 (CADD=22.2, NP_001254479.2:p.Ile26225Thr) in high linkage disequilibrium (LD) with the sentinel SNP rs36051007 (r^2^=0.97, Fig.2b) proposing rs12463674 as the causal variant for HGS at this locus. Notably, we observed prominent clustering and larger HGS effect sizes of ClinVar annotated pathogenic/likely pathogenic PTVs at exons 260-360 encoding for titin’s A-band fibronectin type III-immunoglobulin domains (Fig.2a, TableS6). This is consistent with this region being evolutionally highly conserved and of reported relevance to disease^7^. It further proposes that considering rare and common variant associations jointly may assist clinical variant interpretation.

HGS GWAS loci are enriched for skeletal muscle and central nervous system genes^8^. Consistently, HGS PTV associations were enriched for brain- and muscle-expressed genes, as well as neurodevelopmental and muscle-related pathways (Fig.S5, TableS7). A tight link between brain and muscle function is further supported by our recent finding that PTV-burden in two of the six novel HGS genes, *KDM5B* and *GIGYF1*, is also associated with measures of cognitive function, with *KDM5B* PTVs showing dose-dependent effects in humans and mice [Chen et al., 2022]^9^. Like for cognitive function parameters, *Kdm5b* mutant mice^10^ are characterized by reduced forelimb strength, with heterozygote mice showing intermediate effect sizes to homozygote and wildtype mice, respectively (Fig.3, Fig.S3-S4). We conducted sensitivity analyses correcting HGS PTV-burden associations for educational attainment and reaction time as covariates. Such adjustment had only a minimal impact on the top findings for HGS (TableS8), suggesting that the respective impact of rare variants on cognitive and muscle function is likely related, yet largely independent.

**Figure 3.**
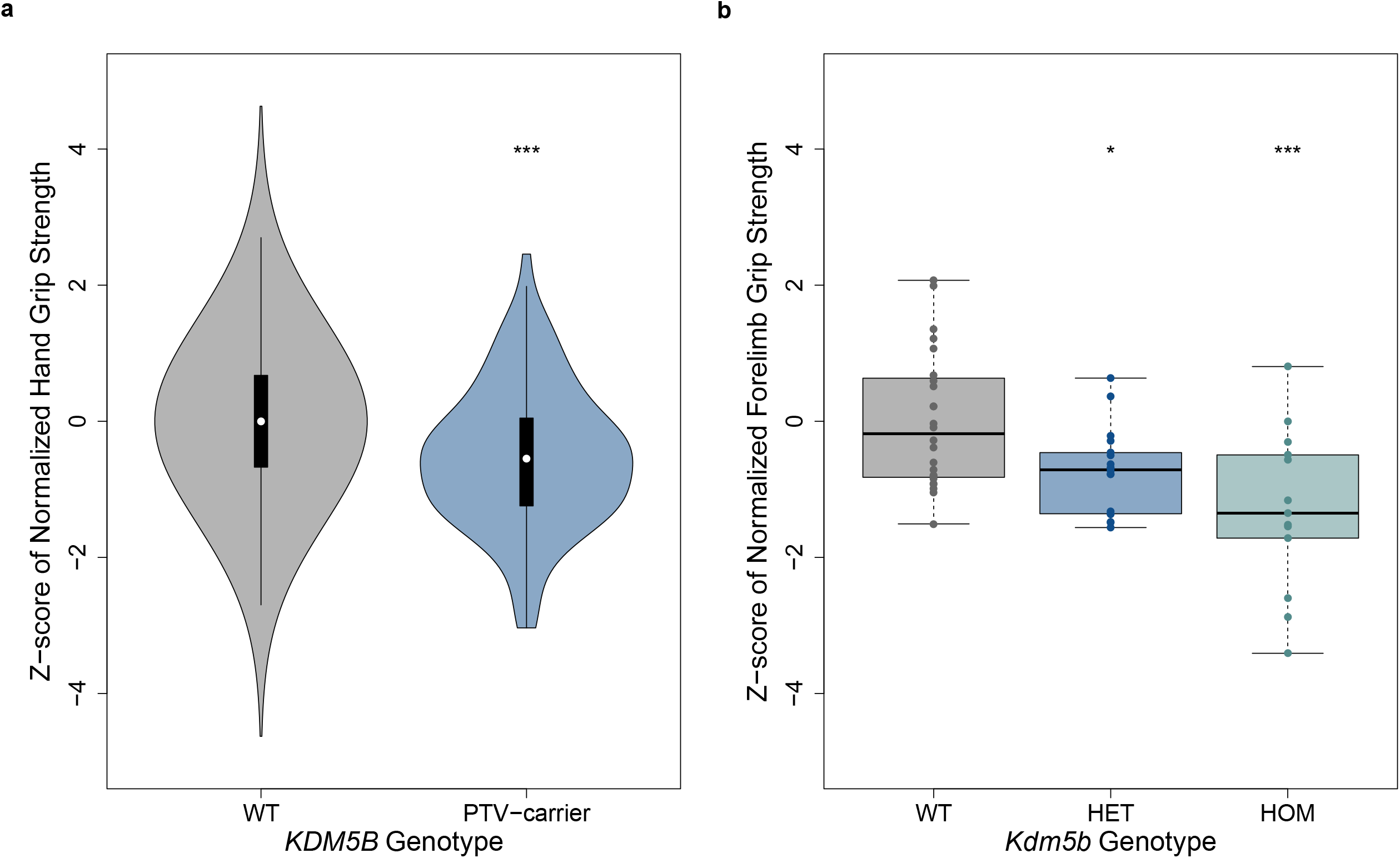
*KDM5B* loss-of-function causes a dose-dependent reduction in HGS in humans and mice. **a)** Normalized HGS stratified for *KDM5B* PTV carrier status. HGS was normalized against standing height, then residualized on age, sex and PCs. Z-scores were generated through inverse-rank normalization, then compared between *KDM5B* PTV carriers vs. non-carriers (WT). **b)** Forelimb grip strength of mice stratified on *Kdm5b* genotype. FGS was normalized against femur length, then corrected for cohort effect. Z-scores calculated from residuals were then compared across heterozygous (HET) and homozygous (HOM) *Kdm5b* mutant and wild type (WT) animals. *p < 0.05; ***p<0.001

Finally, we assessed the interplay between common and rare disease-relevant genetic variation on HGS. A polygenic risk score for HGS (PRS_HGS_) was constructed for 38,118 carriers of PTVs in 199 Mendelian neuromuscular disease (NMD) genes as well as 38,118 randomly selected non-carriers in UKB (TableS9). We observed a significant reduction in HGS for carriers of PTVs in autosomal-dominant NMD genes compared to non-carriers (beta=-0.196kg, p=0.0097), while no reduction was observed for carriers of PTVs in autosomal-recessive NMD genes. Likewise, increase in HGS was significantly associated with per-SD higher PRS_HGS_ (beta=0.954kg, p=1.5e-294). Notably, high PRS_HGS_ mitigated the reduction in HGS for PTV carriers of autosomal-dominant NMD genes, with the effect between PRS_HGS_ and rare PTV-burden in NMD genes being additive (Fig.S6, TableS10).

In summary, we conducted the largest rare-variant association study for muscle strength to date. We identified six genes associated with HGS and support a role for CNS and muscle-related genes and pathways in muscle strength. For *TTN*, we highlight how rare PTVs can assist in the fine-mapping of a GWAS locus and how rare variants ascertained from a biobank population expand the phenotypic spectrum of a medically actionable gene.^7^ Our observation of additive effects between rare coding variation in NMD genes and PRS_HGS_ foreshadow the narrowing gap between common and rare disease genetics and our refined understanding of the genetic basis of human disease resulting from that.

## Data Availability

Summary-level association results produced in the present study are contained in the Supplementary Materials. Additional data produced in the present study are available upon reasonable request to the authors.

## Methods

### The UK Biobank and whole-exome sequencing

The UK Biobank is a large prospective population-based study with over half a million participants recruited across the UK^4^. Phenotypic data collected from each participant includes survey measures, electronic health records, self-reported health information and other biological measurements. The participants have diverse genetic ancestries and overrepresented familial relatedness. Whole-exome sequencing (WES) data from UK Biobank participants was generated by the Regeneron Genetics Center (RGC) as part of a collaboration between AbbVie, Alnylam Pharmaceuticals, AstraZeneca, Biogen, Bristol-Myers Squibb, Pfizer, Regeneron and Takeda. The WES production and quality control (QC) is described in detail in Van Hout et al (2020). As of November 2020, we obtained QC passed WES data (“Goldilocks” set) from 454,787 samples in the UK Biobank.

### Variant annotation

We annotated variants identified through WES by Variant Effect Predictor (VEP) v96 with genome build GRCh38^4^. Variants annotated as stop-gained, splice site disruptive and frameshift variants were further assessed Loss-Of-Function Transcript Effect Estimator (LOFTEE), a VEP plugin. LOFTEE implements a set of filters to remove variants that are unlikely to be disruptive. Those variants labeled as “low-confidence” were filtered out, and we kept variants labeled as “high-confidence”. Variants annotated as missense variants were then annotated by CADD score^11^, which prioritized damaging missense variants. All predicted variants were all mapped to GENCODE canonical transcripts^4^. In total, we identified 714,260 predicted rare PTVs, 6,675,884 missense variants and 3,884,581 synonymous variants with minor allele frequency < 0.1%.

### Phenotyping of hand grip strength

Hand grip strength was measured for both left and right hands using a hydraulic hand dynamometer while seated. Detailed protocol can be found at the UK Biobank data showcase <u>site</u>. Out of 409,559 UK Biobank participants of Caucasian ancestry (Data-Field 22006), we removed individuals with disease diagnoses that can potentially confound hand grip strength measurements including: COPD (N = 2616), brachial plexus disorders (n=50), or history of injuries in elbow, forearm, wrist and hand (n=7608). Due to the large impact of body size on hand grip strength, we restricted our analysis to samples with non-missing and normal body weight (30 to 200 kg).

We also conducted a thorough sensitivity analysis to evaluate which anthropometric covariates to include in order to best capture potential genetic signals not driven-by body size or obesity-related traits. In such analysis, we tested standing height, BMI, as well as whole body fat mass as candidate covariates for hand grip strength GWAS. Adjustment of standing height had a large impact on the number of significant GWAS findings but further adjustment of BMI or whole-body fat mass gave almost identical results. Therefore, we had decided to include standing height as the only anthropometric covariate in our final genetic analysis to avoid unnecessary correction. Samples with grip strength measured for at least one hand was kept in the analysis. The maximum grip strength between two hands was taken as the final outcome for genetic analysis. In total, 340319 samples past QC and filtering were included in the exome analysis.

### Whole-exome and gene-level burden test

We grouped protein coding genes by pLI (v2.1.1)^5^, into LoF intolerant (pLI ≥ 0.9) set and LoF tolerant (pLI < 0.9) set. We annotated rare variants by functional consequences into three types, protein-truncating, missense and synonymous. Missense variants were further annotated by CADD score ^11^ and stratified into 3 groups by predicted deleteriousness, CADD > 30, 30 ≥ CADD > 20, 20 ≥ CADD > 0. In total, we tested hand grip strength association with 15 sets of variants: PTV, CADD > 30, 30 ≥ CADD > 20, 20 ≥ CADD > 0 and synonymous variants in pLI ≥ 0.9 genes; PTV, CADD > 30, 30 ≥ CADD > 20, 20 ≥ CADD > 0 and synonymous variants in pLI < 0.9 genes; and PTV, CADD > 30, 30 ≥ CADD > 20, 20 ≥ CADD > 0 and synonymous variants in all genes. Rare alleles of the same variant category on each gene were aggregated into gene-level burden. The summation of the burden on genes in each gene set was the whole-exome burden.

For the whole-exome burden test, we applied linear regression (“lm” function in R) by fitting whole-exome burden to maximum hand grip strength as the continuous outcome. In the model, we controlled for population structure with top 20 PCs, age, sex, age-squared, age × sex, age-squared × sex, standing height, standing height-squared, standing height × sex, standing height-squared × sex. We also applied the “regenie” ^12^ method to construct genome-wide predictors of hand grip strength using genotype array data and included as model offset to account for additional genetic confounding. We defined a significant threshold at P < 0.0033 (0.05/15) for the whole-exome burden tests.

For gene-level burden test, we fitted linear regression models by regressing hand grip strength on the rare variant burden of each gene. We restricted to PTV and damaging missense (CADD>20) burden for gene-level burden test, based on significant associations in the whole-exome burden tests. We included the same covariates as described above and the corresponding leave-one-chromosome-out (LOCO) predictors constructed by “regenie” ^12^ as model offset. We excluded genes with less than 10 carriers for PTV or damaging missense burden. In total, we tested 15,786 genes for PTV, 17,557 genes for very damaging missense variants (CADD > 30) and 11,370 genes for damaging missense variants (CADD 20-30). Bonferroni correction was applied to declare statistical significance for each category, i.e. p < 3.2e-6 for PTV, p < 2.8e-6 for very damaging missense variants, and p < 4.4e-6 for damaging missense variants.

### GWAS of hand grip strength, MAGMA gene-based analysis and fine-mapping of TTN locus

To assess whether there are converging rare and common variant associations of TTN and OBSCN with hand grip strength, we also conducted a GWAS focusing on common variants with MAF>0.01 in the UK Biobank. 366307 UK Biobank participants of European ancestry post-QC were included in the GWAS of hand grip strength, adjusted for age, sex, age-squared, age × sex, age-squared × sex, standing height, standing height-squared, standing height × sex, standing height-squared × sex, batch effects and top 20 PCs. “regenie” genome-wide predictors were included as model offset to account for additional genetic confounding. A MAGMA gene-based analysis was conducted using FUMA (https://fuma.ctglab.nl/) with the GWAS summary statistics to identify converging common variant associations for *TTN* and *OBSCN*. Statistical finemapping of the *TTN* GWAS locus was conducted using SuSiE^13^ implemented in “susieR” package with default parameters.

### Gene-set PTV-burden analysis

We conducted self-contained gene-set PTV-burden analysis to further capture the impact of rare PTV in particular gene-sets on hand grip strength, including genes with elevated expression in different tissues as well as genes in certain ontology category or biological pathways. Gene-set PTV-burden is constructed by summing the rare alleles of PTVs of all genes in each gene-set, association testing was carried out using linear regression (“lm” function in R) by fitting gene-set burden to maximum hand grip strength as the continuous outcome. In the model, we controlled for population structure with top 20 PCs, age, sex, age-squared, age × sex, age-squared × sex, standing height, standing height-squared, standing height × sex, standing height-squared × sex. We also applied the “regenie” ^12^ method to construct genome-wide predictors of hand grip strength using genotype array data and included as model offset to account for additional genetic confounding.

#### Genes with elevated expression in different tissues

We explored how rare PTV-burden of genes enriched in different tissues impact hand grip strength based on data from the Human Protein Atlas (HPA, http://www.proteinatlas.org). Based on data from the HPA, we extracted 10992 genes that showed elevated expression in at least 1 of 36 different tissues including a range of 51 genes for smooth muscle to 2709 genes for brain. We calculated PTV-burden for genes that showed elevated expression in each tissue. Rare alleles of the PTV on each gene were aggregated into gene-level burden; the summation of the burden on genes was the gene-set burden. A false discovery rate < 0.05 for 36 tissues was applied to declare significance for this analysis.

#### Pathway-based gene-set PTV-burden

We also constructed pathway-based gene-set PTV-burden for 20928 gene-sets from MSigDB v7.2 ^14^ including 10266 Gene Ontology (GO) terms (7569 biological processes, 1001 cellular components, 1696 molecular functions), 4493 Human Phenotype Ontology (HPO) terms, 3354 curated chemical and genetic perturbation (CGP) terms, and 2815 curated canonical pathways from BioCarta, KEGG, PID, Reactome and WikiPathways.^14^ Rare alleles of the PTV on each gene were aggregated into gene-level burden; the summation of the burden on genes in each pathway-based gene-set was the gene-set PTV-burden. The pathway-based PTV-burden association with hand grip strength was assessed in a self-contained manner and a false discovery rate < 0.05 was used to declare statistical significance. (Table S11)

### Phenome-wide association study (PheWAS)

We performed a PTV-burden of *TTN* PheWAS across 3,654 binary and 238 quantitative phenotypes. Each binary phenotype was either derived from an ICD10 diagnosis code in the UK Biobank and mapped to a Phecode, or derived from self-reported illnesses, operation procedures or medication use. We excluded phenotypes with less than 100 cases for binary phenotypes. We took a two-step approach to first test all gene-phenotype pairs by logistic regression and then performed Firth’s logistic regression for those gene-phenotype pairs passed significant threshold (P < 0.01). For quantitative phenotypes, we excluded phenotypes with fewer than 100 observations and phenotypes with less than 12 distinct values. For each phenotype, we removed individuals with phenotype value > 5 SDs from the sample mean. Burden testing was performed using linear regression on both the raw and inverse rank-based normal transformed quantitative phenotypes. We controlled for population structure with top 20 PCs, age, sex, age-squared, age × sex, age-squared × sex in the PheWAS. We defined phenome-wide significant thresholds as P < 1.3e-5 (0.05/3892).

### Polygenic risk score (PRS) analysis

To assess the interplay between common variants and rare PTVs for hereditary neuromuscular disease genes on hand grip strength, we first developed a PRS for hand grip strength in the UK Biobank Caucasian samples excluding 38118 carriers of PTVs in 199 genes associated with Mendelian NMDs as well as 38118 randomly selected non-carriers. A GWAS of hand grip strength was then conducted using the rest of the UK Biobank samples in the same manner as described above. A PRS for hand grip strength was then constructed based on the summary statistics using the PRS-CS method^15^ with default settings and no pre-specified phi value. LD reference panel was precomputed using 1000 Genomes Project phase 3 samples with European ancestry (available at https://github.com/getian107/PRScs). PRS of each chromosome for each individual in the validation set was computed by the “--score” function in PLINK 2.00 alpha, a linear combination of genotypes weighted by effect size estimates. The final PRS was then summed across chromosome 1 to 22. We then tested the contribution of hand grip strength PRS and PTV-burden in Mendelian NMD genes on hand grip strength in the UKBB reserved samples (38118 carriers of PTVs in 199 genes associated with Mendelian NMDs as well as 38118 randomly selected non-carriers), as well as the interaction between PRS and PTV-burden. We binarized PTV-burden to carriers vs. non-carriers. To have a better understanding of the potentially different contributions of PTVs in Mendelian NMD genes with dominant vs. recessive inheritance, we further categorized PTV-carriers into carriers for autosomal dominant NMDs vs. autosomal recessive NMDs. (Supplementary Table S9) We then applied linear regression with hand grip strength as a continuous outcome:

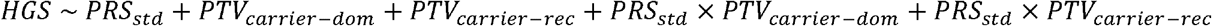

HGS: hand grip strength (maximum); PRS_std_: standardized hand grip strength PRS;

PTV_carrier-dom_: PTV carriers for genes of autosomal dominant Mendelian NMDs;

PTV_carrier-rec_: PTV carriers for genes of autosomal recessive Mendelian NMDs;

In the model, we also controlled for population structure with top 20 PCs, age, sex, age-squared, age × sex, age-squared × sex, standing height, standing height-squared, standing height × sex, standing height-squared × sex.

### In vivo KDM5B experiment

#### Animals

The generation of a mouse Kdm5b loss of function allele (MGI:6153378), its breeding and housing has been previously reported^10^ [Chia-Yen et al.,2022]. Briefly, generation was through a CRISPR/CAS9 mediated deletion of coding exon 7 (ENSMUSE00001331577), leading to a premature translational termination due to a downstream frameshifted reading frame. Breeding of test cohorts was performed on a C57BL/6NJ background. Breeding, housing, and all experimental procedures with mice were approved by the Animal Welfare and Ethical Review Body of the Wellcome Sanger Institute and, conducted under the regulation of UK Home Office license (P6320B89B), and in accordance with institutional guidelines. At 14-15 weeks of age, 24 wildtype, 18 heterozygous and 13 homozygous Kdm5b mutant male mice were tested, in two independent cohorts.

#### Grip Strength Measurement

A Grip Strength Meter (BIO-GT3+MR; Bioseb, France) with a custom grid attachment (stainless steel, 9.8 × 11cm, with 2mm bars 0.85cm apart, German Mouse Clinic, Neuherberg, Germany) was used to measure forelimb grip strength. Experimenters were blind to genotype. As a mouse grasped the grid with the forepaws, it was pulled off the grid and the peak pull force in grams was recorded on a digital transducer. This was repeated three times for each mouse and the mean value was used. Grip strength normalisation to body size was calculated as the mean grip strength divided by femur length.

#### X-rays

Half the mice were anaesthetised with ketamine/xylazine (100mg/10mg per kg of body weight) and then placed in an MX-20 X-ray machine (Faxitron X-Ray LLC). Whole body x-radiographs were taken in a dorso-ventral orientation. A second set of x-radiographs were generated from independent animals using post-mortem dissected legs. All images were analysed and morphological abnormalities assessed using Sante DICOM Viewer v7.2.1 (Santesoft LTD).

#### Statistical analysis (mouse data)

All statistical analyses of mouse data were performed using R. Data was first transformed to achieve normality, using Box Cox transformation (MASS package, with lambda limit = [-2,2]). Testing for genotype effect was performed using a double generalised linear model, dglm (dglm package). Cohort was used as a covariate for the Box Cox transformation as well as the dglm test. For visualization purposes, residual values were calculated from the linear model and z-scores relative to wildtypes were calculated.

