## Supplementary Information for "Rare genetic variants impact muscle strength"

### Discussion of significant genes from gene-level PTV-burden association analysis of hand grip strength

We conducted gene-level PTV and missense-burden association analyses to identify individual genes in which a burden of rare coding variants affect hand grip strength. In total, 15,786 genes with at least 10 carriers of PTVs with MAF<0.001 were included in the gene-level burden analysis. We found 6 genes (*KDM5B*, *OBSCN*, *GIGYF1*, *TTN*, *RB1CC1* and *EIF3J*) for which PTV-burden showed significant association with hand grip strength after Bonferroni correction ($p < 3.2e-6$). Seven additional genes showed a false discovery rate (FDR)<0.05 (Fig.1b, TableS2). We did not identify missense-burden associations at the level of individual genes (TableS3-S4).

*TTN* was discussed in the main text.

*OBSCN*

Obscurin is another giant sarcomeric protein. It is the third member of the sarcomeric giant protein family that, along with titin and nebulin, represents the underlying backbone that guides the assembly of the sarcomere and maintains its stability over time in striated muscle. In adult striated muscles, obscurin shows a definitive localization pattern with a larger distribution at the M-bands, bridge titin and myomesin at the M-band. [1,2] Homozygous LoF variants in *OBSCN* were recently identified to cause severe recurrent rhabdomyolysis[3]. Intriguingly, we found a higher grip strength for 3,834 heterozygous *OBSCN* PTV-carriers compared to non-carriers of UKB (beta=0.69kg, p=4.4e-10). No reduction in HGS was observed for 470 heterozygous carriers of the pathogenic LoF variants reported to cause severe recurrent rhabdomyolysis[3] (Figure S2).

*KDM5B*

Lysine-specific demethylase 5B (KDM5B, also called JARID1B or PLU1) is a JmjC domain containing enzyme that removes methyl groups from tri- or di-methylated H3K4 (H3K4me3/2) to produce monomethylated H3K4 (H3K4me1).[4] KDM5B is overexpressed in breast-, prostate-, bladder-, lung- and melanoma cancer, and serves as a potential oncogene. KDM5B has been shown to interact with the HDAC complexes. It has been shown, that HDAC inhibition decreased the level of KDM5B expression.[5] Histone deacetylases (HDACs), which targets histone and non-histone proteins, is a major enzyme family that controls the biological process of histone deacetylation, hence HDACs are essential for gene expression, metabolic and physiological function of the skeletal muscle system. Recent studies have shed light on the role of HDACs in skeletal muscle metabolism, myogenesis, maintaining sarcomere homeostasis, mitochondria remodeling, etc.[6] KDM5B also plays an important role in early embryonic development and mutations in *KDM5B* lead to development delay and intellectual disability.[7] Given the correlation observed between hand grip strength and cognitive function[8] it is likely that part of the association between *KDM5B* PTV-burden and hand grip strength can be explained by cognitive deficits. However, our sensitivity analysis controlling for baseline education level or reaction time had minimal impact on the association signal suggesting other uncaptured developmental mechanisms through which the brain-related genes might impact muscle strength.

*GIGYF1*

*GIGYF1* encoding for GRB10-interacting GYF protein 1 can modulate insulin-like growth factor 1 receptor (*IGF-1R*) signaling pathway.[9] Upon the binding of IGF-1 to the IGF-1R, several important signaling pathways get activated, such as the PI3K/Akt-, the Akt/mTOR- and the GSK3β pathways. There is also a crosstalk between IGF-1 and myostatin signaling pathways. The IGF-1 and its receptor are key regulators of both anabolic and catabolic pathways.[10] It has been recently reported that loss-of-function variants in *GIGYF1* associated with risk of type II diabetes and increased glucose levels.[11]

*RB1CC1*

Rb1-inducible coiled-coil 1 (Rb1cc1) is a DNA-binding protein and is abundantly expressed in human musculoskeletal cells.[12] Rb1cc1 expressed at high levels is associated with the maturation of human embryonic musculoskeletal cells, thus, Rb1cc1 is prerequisite for myogenic differentiation.[13]

### *EIF3J:*

EIF3J encodes for eukaryotic translation initiation factor 3 subunit J, which is a core subunit of the eukaryotic initiation factor 3 complex participating in the initiation of translation by aiding in the recruitment of protein and mRNA components to the 40S ribosome. (RefSeq, Sep 2013) It is highly expressed in skeletal muscle[14] but its potential impact on muscle function is not clear.

**Figure S1.** Phenome-wide association of TTN PTV-burden in the UK Biobank. 3,654 binary and 238 quantitative phenotypes were tested, red dashed line indicates the Bonferroni-corrected significance threshold. Top nine associations were annotated.

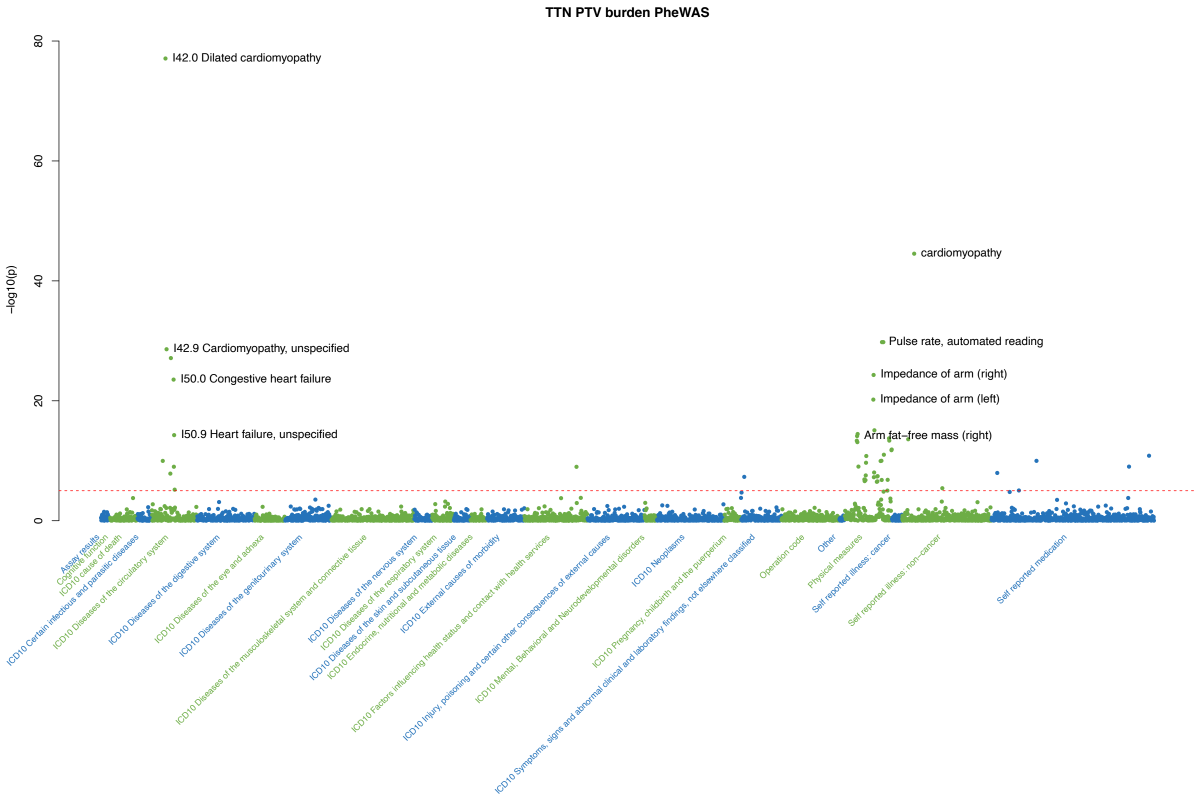

**Figure S2.** Single PTV association of OBSCN with HGS in the UK Biobank. Effect sizes of each PTV in OBSCN were plotted against its genomic position, with exon numbers and protein domains demonstrated at the bottom. ClinVar pathogenic/likely pathogenic PTVs were colored as red diamond. Pathogenic LoF variants recently reported for severe recurrent rhabdomyolysis[3] were highlighted in dark red.

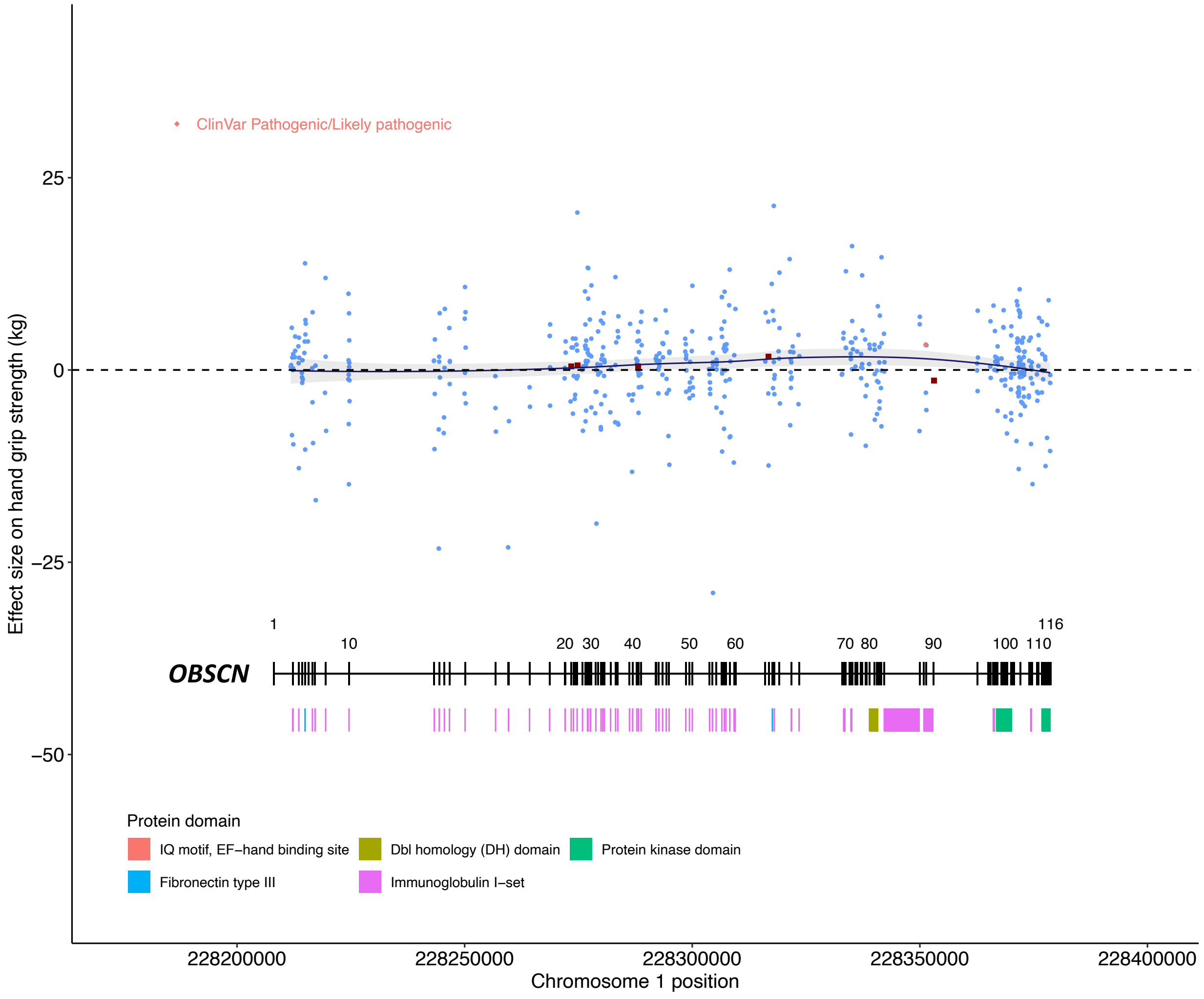

**Figure S3.** Forelimb grip strength (FGS, unnormalized) of heterozygous (HET) and homozygous (HOM) Kdm5b mutant mice vs. wild type (WT). FGS was corrected for cohort effects. Z-scores calculated from residuals were compared across different genotypes.

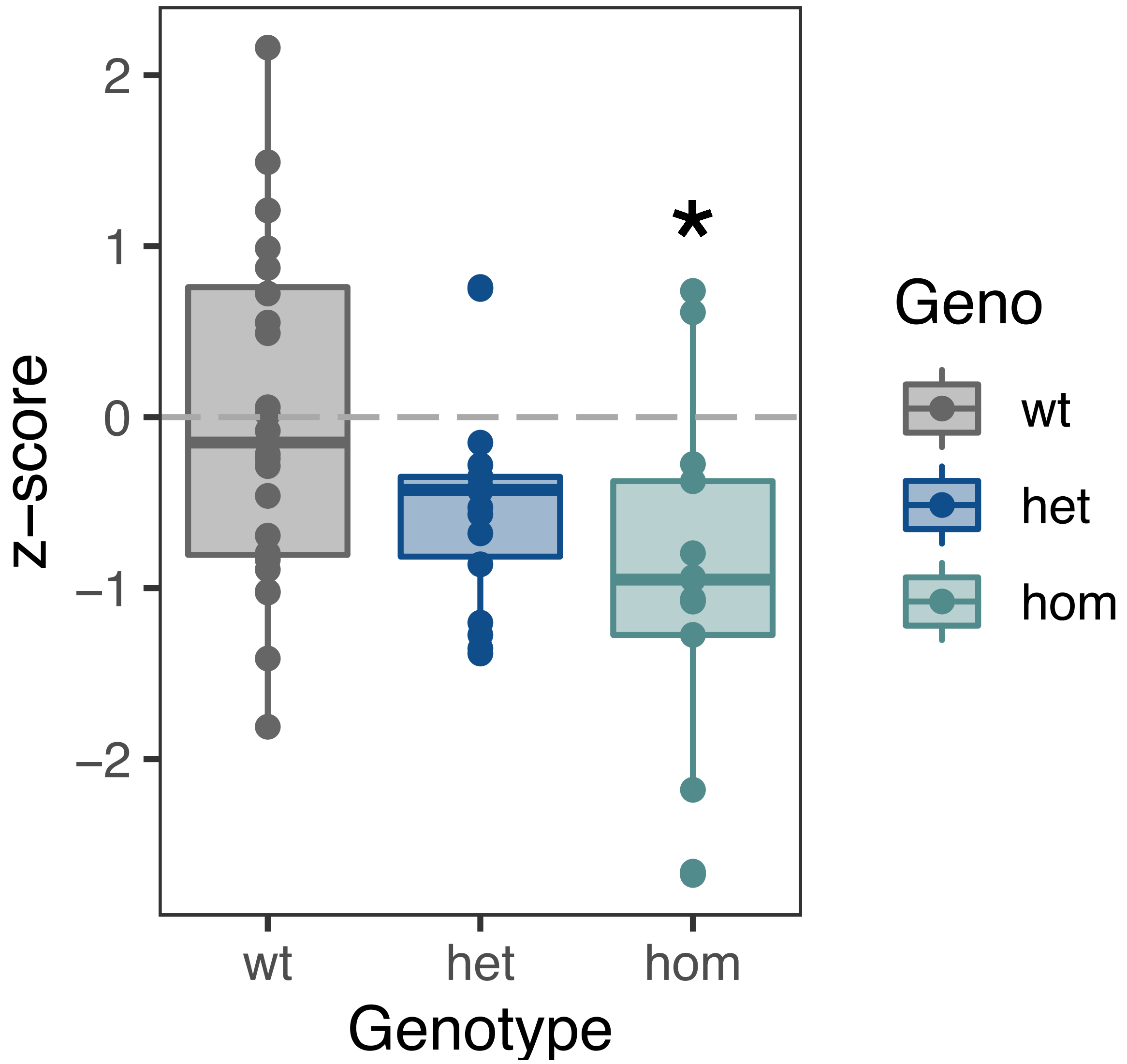

**Figure S4.** Correlation of hand grip strength with standing height in humans and forelimb grip strength with femur length in mice. Simple linear regression line was plotted. Pearson's correlation coefficient and p-value were shown.

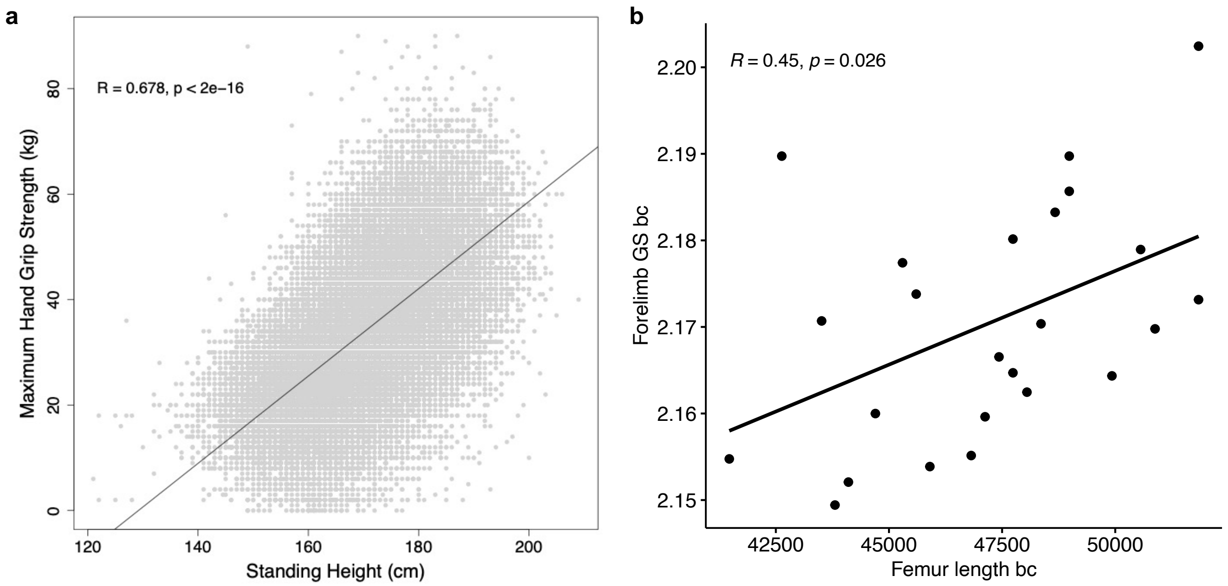

**Figure S5.** Tissue-expressed gene-set PTV-burden associations with hand grip strength. Gene sets were constructed based on 10992 genes that showed elevated expression in at least 1 of 36 different tissues from Human Protein Atlas. Number of genes, effect size of PTV-burden on hand grip strength and false discovery rate were shown for each tissue-expressed gene set.

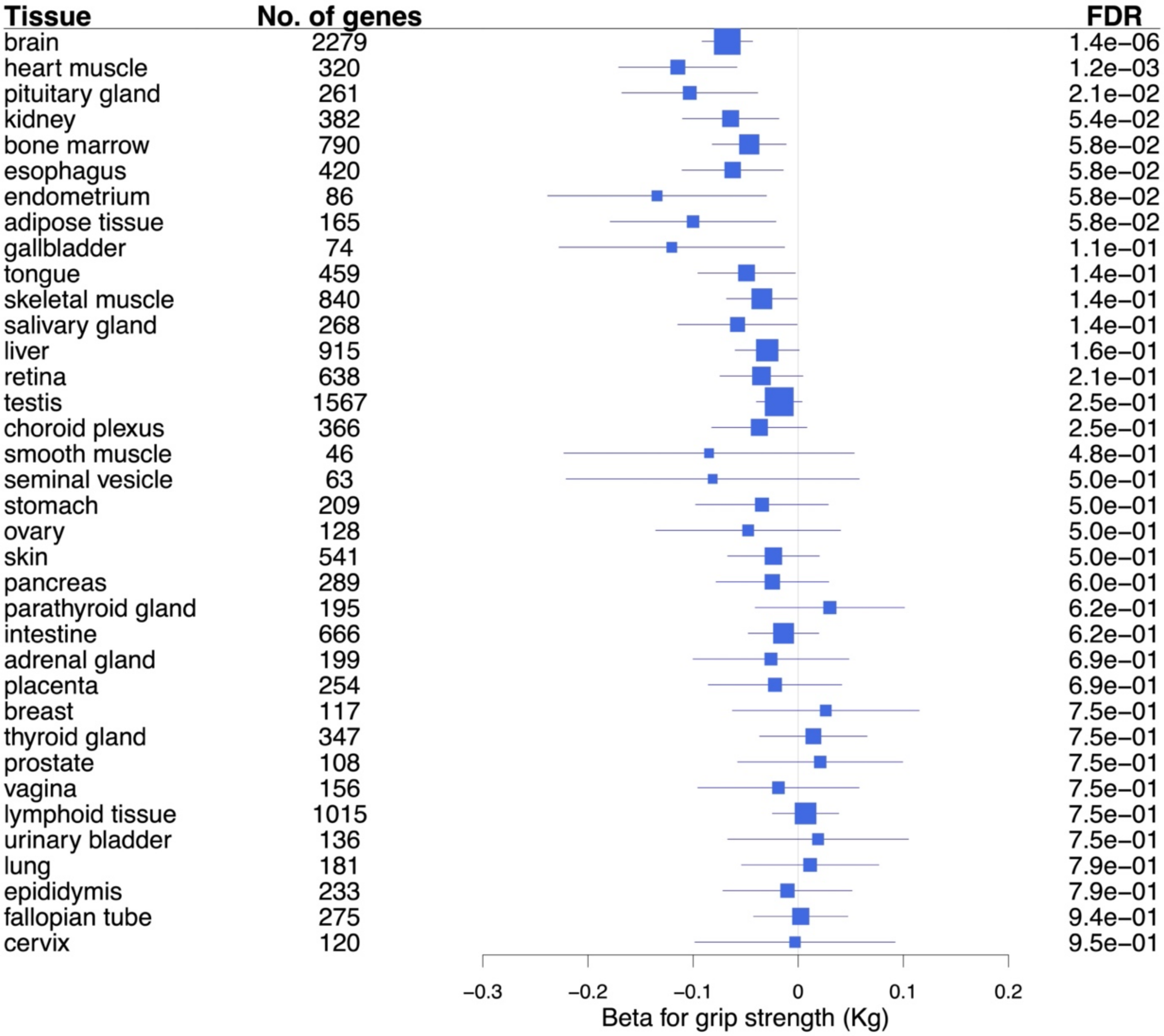

**Figure S6.** Polygenic risk score (PRS) effects on hand grip strength (HGS) stratified for PTV carrier status of Mendelian neuromuscular diseases (NMDs). Mean residualized HGS was plotted against percentiles of HGS-PRS for carriers of PTVs in autosomal dominant NMD genes, carriers of PTVs in autosomal recessive NMD genes, and non-carriers.

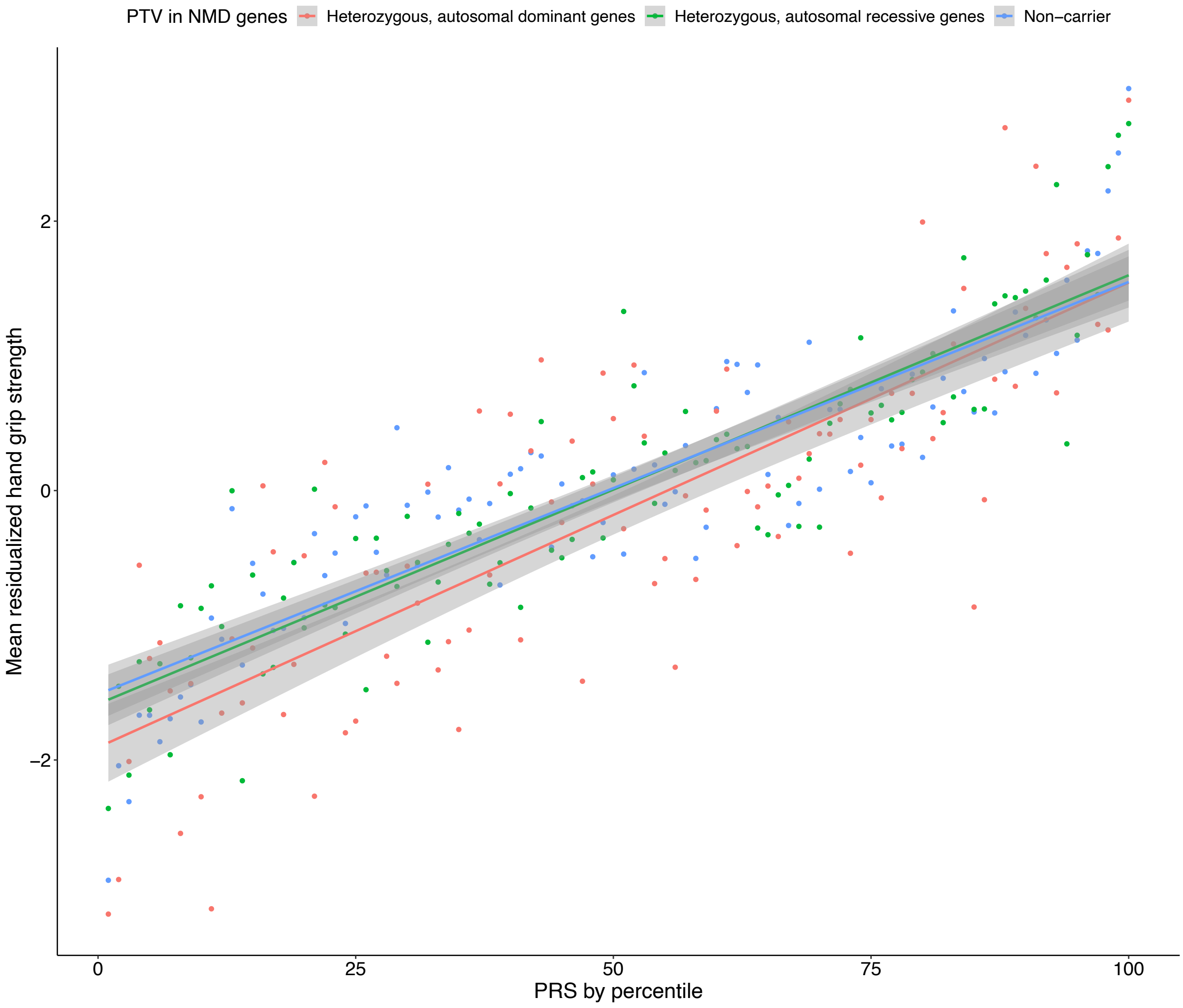
